# Adjusted fatality rates of COVID19 pandemic: a comparison across countries

**DOI:** 10.1101/2020.05.13.20099796

**Authors:** Carlos Canelo-Aybar, Jessica Beltran, Marilina Santero, Pablo Alonso-Coello

**Author notes:** **Corresponding author, *Carlos Canelo-Aybar***, Iberoamerican Cochrane Centre - Department of Clinical Epidemiology and Public Health, Biomedical Research Institute Sant Pau (IIB Sant Pau), Sant Antonio María Claret 167, 08025 Barcelona, Spain Telf.

## Abstract

**Background:** A key impact measure of COVID-19 pandemic is the case fatality rate (CFR), but estimating it during an epidemic is challenging as the true number of cases may remain elusive.

**Objective:** To estimate the CFR applying a delay-adjusted method across countries, exploring differences to simple methods and potential correlation to country level variables.

**Methods:** Secondary analysis of publicly available data from countries with ≥500 cases by April 30^th^. We calculated CFR adjusting for delay time from diagnosis to death and using simple methods for comparison. We performed a random effects meta-analysis to pooling CFRs for all countries and for those with high testing coverage and low positivity rate. We explored correlation of adjusted CFR with age structure and health care resources at country level.

**Results:** We included 107 countries and the Diamond Princess cruise-ship. The overall delay adjusted CFR was 2.8% (95%CI: 2.1 to 3.1) while naive CFR was 5.1% (95%CI: 4.1 to 6.2). In countries with high testing coverage/low positivity rate the pooled adjusted CFR was 2.1% (95%CI: 1.5 to 3.0), there was a correlation with age over 65 years (β = 0.12; 95%CI: 0.06 to 0.18), but not with number of physician or critical care beds. Naive method underestimated the CFR of the CFR with a median of 1.3% across countries.

**Conclusion:** Our best estimation of CFR across countries is 2% and varies according to the aged population size. Modelers and policy makers may consider these results to assess the impact of lockdowns or other mitigation policies.

## 1. Introduction

The coronavirus disease 2019 (COVID-19) is an infectious illness caused by the severe acute respiratory syndrome coronavirus 2 (SARS-CoV-2), which was first reported in December 2009 in Wuhan, China. The disease has since then spread rapidly through mainland China and worldwide [1]. On March 12th 2020, the COVID-19 outbreak was officially characterized as a pandemic by the World Health Organization (WHO) [2]. As of April 30th, globally, nearly three million confirmed infections have been reported, together with more than 250,000 deaths due to COVID-19 [3]. Travel restrictions and lockdowns have been imposed by many countries as actions to mitigate the epidemic growth, with around three billion people living in in countries with border constraints [4], and countries like India, China, France, Italy, New Zealand, Poland, UK, and Spain have implemented restrictive mass quarantines [5].

During an outbreak, case fatality rate (CFR) provides an estimate of the magnitude of an infectious disease, frequently calculated by dividing cumulative deaths by cumulative cases at any moment (naive method). This estimate should be distinguished from the infectious fatality rate (IFR), which is calculated over the total number of infected people (usually estimated from seroprevalence studies). However, the naive approach is subject to several biases in different directions, including preferential ascertainment of severe cases and reporting delay bias [6]. Ascertainment bias occurs when there are asymptomatic cases or with mild symptoms who are not tested and not included in the denominator, while a delayed reporting bias is due to a tome lag between the moment a case is confirmed to the time of death. This is specially concerning when there is a rapidly increasing incidence [6]. Due to these factors, as the epidemic progresses, and cases are tracked more appropriately, naive estimates may falsely suggest a rise in the CFR, which can increase the public alarm [7, 8].

As an example, of the limitations described above, during the current COVID-19 pandemic the WHO initially reported a CFR of 2% in a press conference on January 29; this estimate was subsequently updated up to 3.4% on March 3, suggesting and increase trend on the naive estimate. These calculations were further limited by the fact that the different CFRs should not directly be averaged across countries, as each country will likely have a different testing coverage, age structure or healthcare capacity, all factors that will have an impact in the number of fatalities.

Given the relevance of obtaining a reliable estimation of the fatality rate during the COVID-19 outbreak, we aim to aim to provide more accurate estimations of the COVID-19 CFR across countries using previously published methods(7), and standard pooling meta-analysis techniques. Finally, we will also explore the potential correlation to country level variables.

## 2. Material and Methods

### 2.1 Data sources

This is a secondary analysis of public available data from several sources including the situation reports prepared by WHO [9], the John Hopkins University interactive track map (https://coronavirus.jhu.edu/map.html) which compiles data from several public sources [10] and the Our World in Data repository which also contains testing statistics (https://ourworldindata.org/coronavirus) [11]. We obtained data about population size and proportion over 65 years old from the World Bank demographics statistics [12], the number of critical care beds from previous published studies[13, 14], and the number of healthcare workers from the World Health Organization observatory [15].

### 2.2 Inclusion criteria

We considered data from countries with at least 500 of reported cases with the SARS-Cov-2 by April 30th. This threshold was based in our sample size calculation to estimate a single population proportion of 3% with a 95% confidence interval and a margin of error of +/− 1.5% [16].

### 2.3 Fatality rate estimates

We collected data per country on the total number of confirmed cases, total number of cases recovered, and total number of deaths from January 22 to April 30th. Based on these three variables we estimated the CFR using the following methods:

I. We estimated the *delay adjusted CFR (dCFR) applying* a correction method previously described by Nishiura et al, which accounts for the delay from case confirmation to death or recovery [17]. This method has recently been used to correct the CFR from the Diamond Princess cruise ship [18], and the programing code was made publicly available by the authors (https://github.com/thimotei/cCFRDiamondPrincess) based in the following formula [18]:

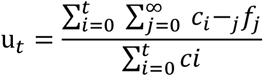 Where *u_t_* is the correction factor applied to the denominator (cumulative number of cases), to estimate the proportion with known outcome, *c* is the daily case incidence at time *i and* f is the proportion of cases with delay between onset or hospitalization and death [18]. To apply this method we assumed a similar time distribution from hospitalization (which we assumed to be equivalent to the date of diagnosis in our analysis) to death, with a mean of 13 days, based on an analysis of 39 cases of COVID-19 from the city of Wuhan, China [19].
II. For comparison purposes we the *naive CFR (nCFR)*, which was obtained by dividing the cumulative number estimated of deaths by the cumulative number of cases; and the *recovery CFR (rCFR)* obtained by dividing the cumulative number of deaths by the cumulative number of deaths and recovered patients.

### 2.4 Data Analysis

To pool the CFR across countries, we performed a meta-analysis of proportions using a generalized linear mixed random model (which assumes a distribution of different true estimates across entities) with a logit transformation, and the Clopper Pearson method to estimate the confidence intervals for each observation [20]. We assessed the heterogeneity in the estimated CFR across countries by visual inspections of the forest plots, as the Q statistic and the I-square parameter are not recommended for to evaluate inconsistency for proportion meta-analysis. We performed the analysis using the package *metaprop* in the R statistical platform, version 3.6.2.

We provided a pooled delay adjusted CFR estimate for countries with a good coverage of COVID-19 testing, which we defined for purpose of our analysis as having performed more than 10 tests per thousand habitants and maintaining a rate of positive tests lower than 10%, and as a sensitivity analysis we performed the same analysis for countries with less than 5% of positivity rate. Countries with these characteristics are more likely able to maintain an acceptable surveillance of positive cases as the outbreak progresses and, thus typically provide more reliable CFR estimates.

Within this group of latter group of countries, we also conducted an exploratory ecological analysis to assess if there were a positive correlation between the dCFR and the proportion of population over 65 years old, or a negative correlation with the number of critical care beds and the number of physicians. We performed three independent bivariate linear regression analysis after a logarithmic transformation of the delay adjusted CFRs, and a multivariate regression analysis including variables with a *p value* <0.10. We tested the normality of residuals by producing a kernel density plot and homoscedasticity by plotting residuals versus fitted (predicted) values.

Finally, we graphically plotted the progression of the nCFR and the dCFR over time, since the seventh day after the first reported death, for three countries that showed a flattened epidemic curve over time (New Zealand, South Korea, and Germany).

## 3. Results

We included data from a total of 107 countries with more than 500 confirmed cases reported, most of them from Europe (39), and Asia (32). We also included data from the 712 cases among passengers of the Diamond Princess cruise ship. As of April 30^th^, the number of cumulative cases reported ranged from 539 (Georgia) to 1,069,424 (United States), while the cumulative number of deaths reported ranged from 2 (Djibouti) to 62,996 (United States). The countries with the largest proportion of population over 65 years old were Japan and Italy; the countries with the largest number of critical beds per habitants were the United States, Germany and Turkey; and the countries with the largest number of physicians were Cuba, Sweden and Austria. **(*Supplementary Table S1*)**.

We identified a subset of 26 countries, together with the Diamond Princess cohort as having large number of tests performed and low positivity rate. Among these countries, the one with the highest number of tests performed was Iceland (120.4 tests per thousand people), and the lowest was South Korea (10.7 tests per thousand people). The highest positivity rate was observed in Singapore (9.7%) and the lowest in Hong Kong (0.8%).

The dCFR ranged from 0.2% (Qatar) to 24% (United Kingdom), our overall pooled estimation showed a dCFR across countries of 4.3% (95%CI: 3.5 to 5.1). There was important heterogeneity, with a group of 12 countries showing a CFR above 15%, most of them from the European region apart from Mexico (20.9%), Honduras (15.3%) and Brazil (15.2%). The overall pooled naive estimates underestimated the CFR by 35%, showing a rate of 2.8% (95%CI: 2.3 to 3.3) across countries. The difference between the nCFR and the dCFR across countries had a median of 1.3% and ranged between 0.1% (Hong Kong, Iceland, China) to than 12% (Mexico), being correlated to the intensity of the outbreak progression. The *recovery method* showed a much higher fatality rate (CFR 16.0%; 95%CI 11.9% to 21.3%), but many of the individual values per country were implausible (over 50%), precluding the possibility of further analysis using this method ***(Supplementary Figure S1)***.

In the subgroup with large testing coverage, the dCFR ranged from 0.2% (Singapore) to 9.6% (Canada). Our initial pooled estimation was 2.3% (95% CI 1.6% to 3.3%) ***(Supplementary file figure S2)***, but at the visual assessment of the forest plot there was important heterogeneity with the largest values for countries like Denmark (6.6%), Slovenia (7.4) and in particular for Canada (9.6%). The dCFR from Canada was excluded in a subsequent analysis because their confidence interval did not overlap with any the estimates of the remaining countries, this analysis showed a pooled dCFR of 2.1% (95%CI 1.5 to 3.0), and a prediction interval that ranged from 0.4% to 10.9% (**Figure 2)**. Our sensitivity analysis including countries with positivity rate ≤5% showed a consistent result with a pooled dCFR of 2.0% (95%CI 1.5 to 2.8) ***(Supplementary file figure S3)***.

**Figure 1.**
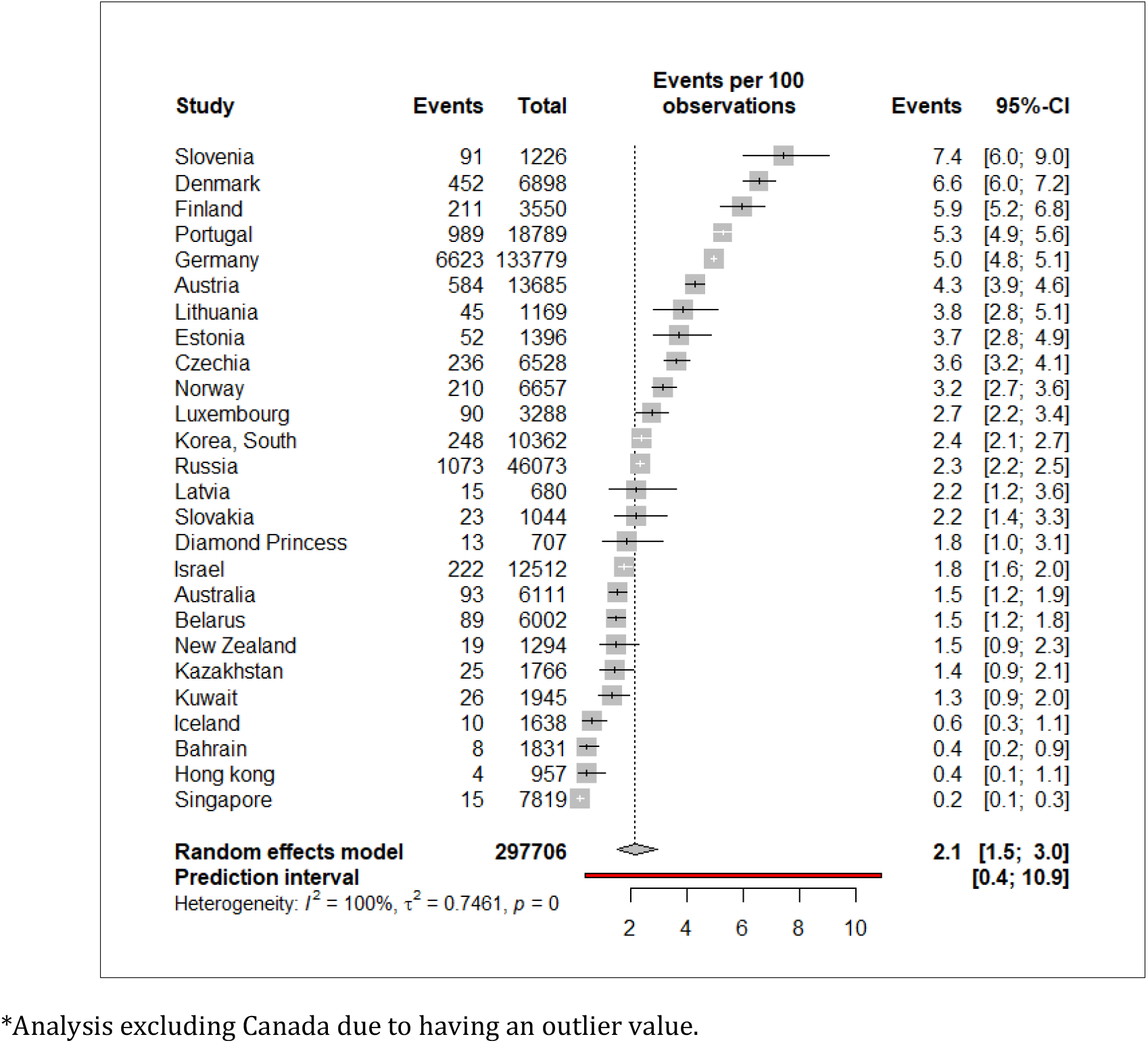
Meta-analysis of the delay adjusted CFR across countries with high testing coverage and low positivity rate

**Figure 2.**
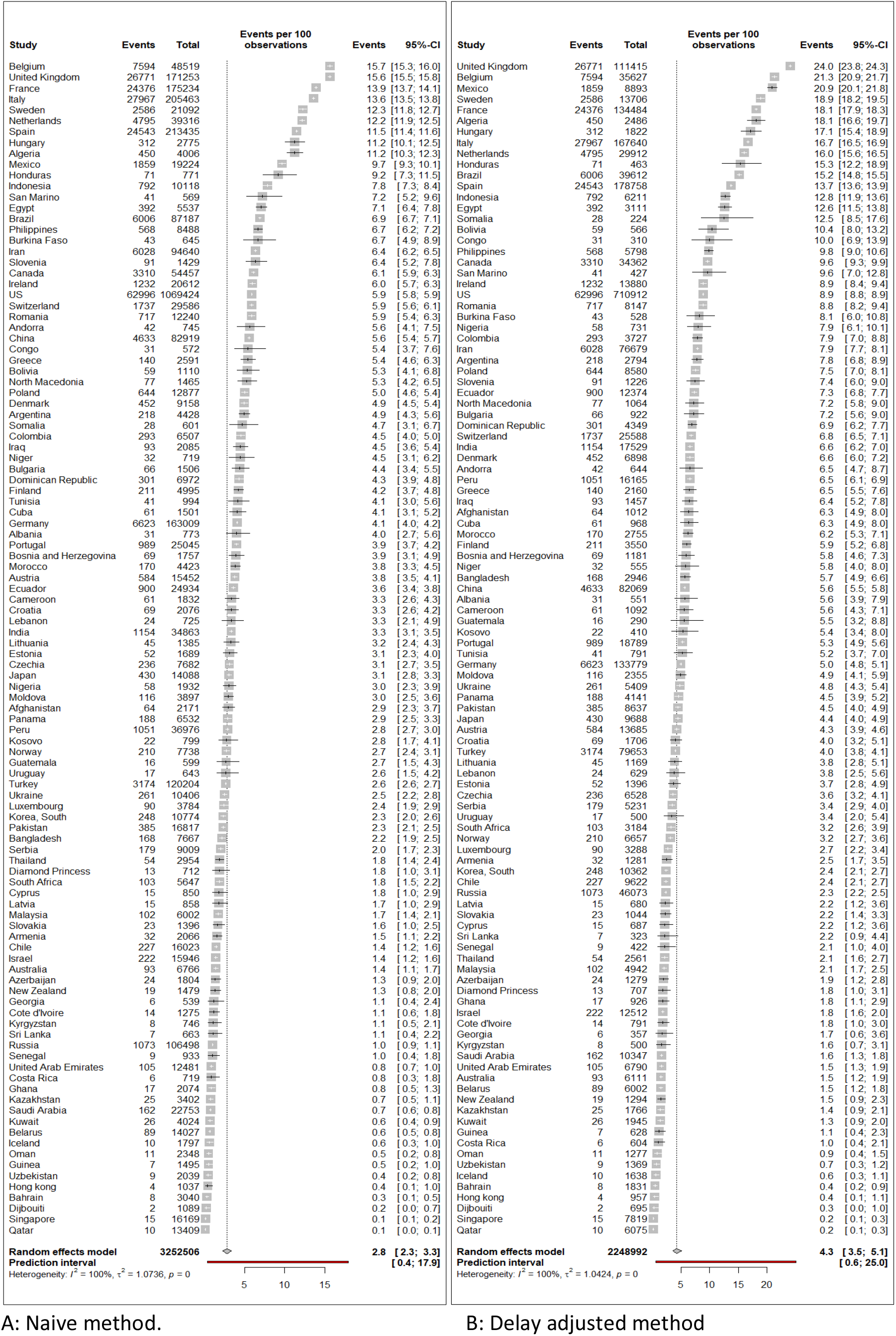
Overall meta-analysis across countries of the naive and delay adjusted CFR.

The linear regression analysis among the countries with larger testing coverage did not show an association between the dCFR and the number of critical care beds per million people (β = 0.01; 95%CI = −0.07 to 0.08, *p value=* 0.85); but we observed and association with the number of physicians per thousand people (β = 0.52; 95%CI = 0.18 to 0.87, *p* value= 0.004) and with the proportion of population over 65 years old (β = 0.11; 95% CI 0.06 to 0.18, *p value* <0.001) (**Figure 3**). In the multivariate analysis, there was no association with the number of physician (β = 0.26; 95%CI = −0.09 to 0.63, *p value* 0.14), while the association with age over 65 years remained statistically significant (β = 0.09; 95%CI = 0.02 to 0.16, *p value=* 0.01).

**Figure 3.**
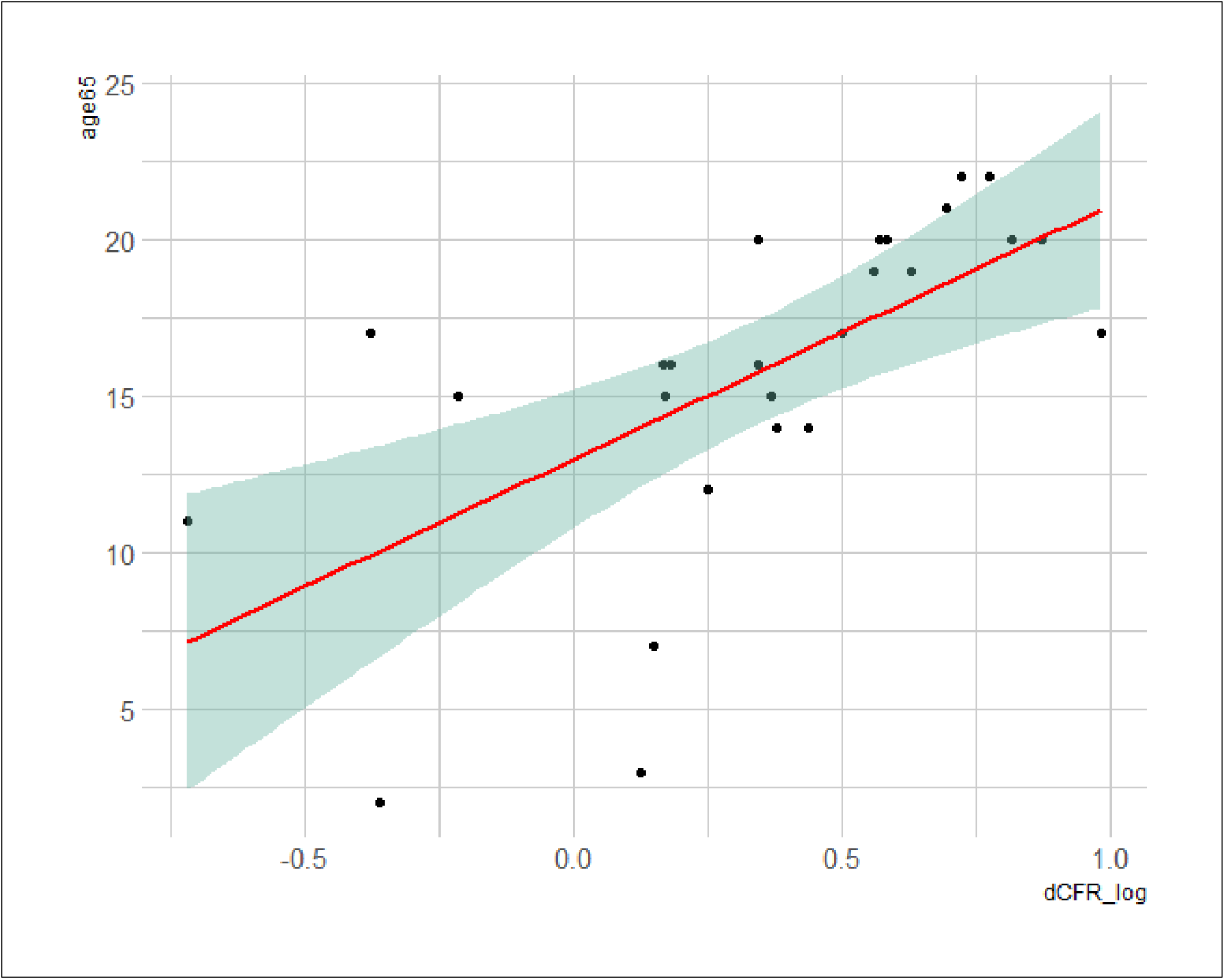
Linear regression plot for proportion of population over 65 years and the delay adjusted CFR.

We plotted the nCFR and dCFR over time for New Zealand, South Korea, and Germany. In the three examples, we observed that, as the epidemic progressed over time the naive estimations showed an increasing pattern, from very low rates to rates near the adjusted estimations, with a lag of approximately one to two weeks (**Figure 4**).

**Figure 4.**
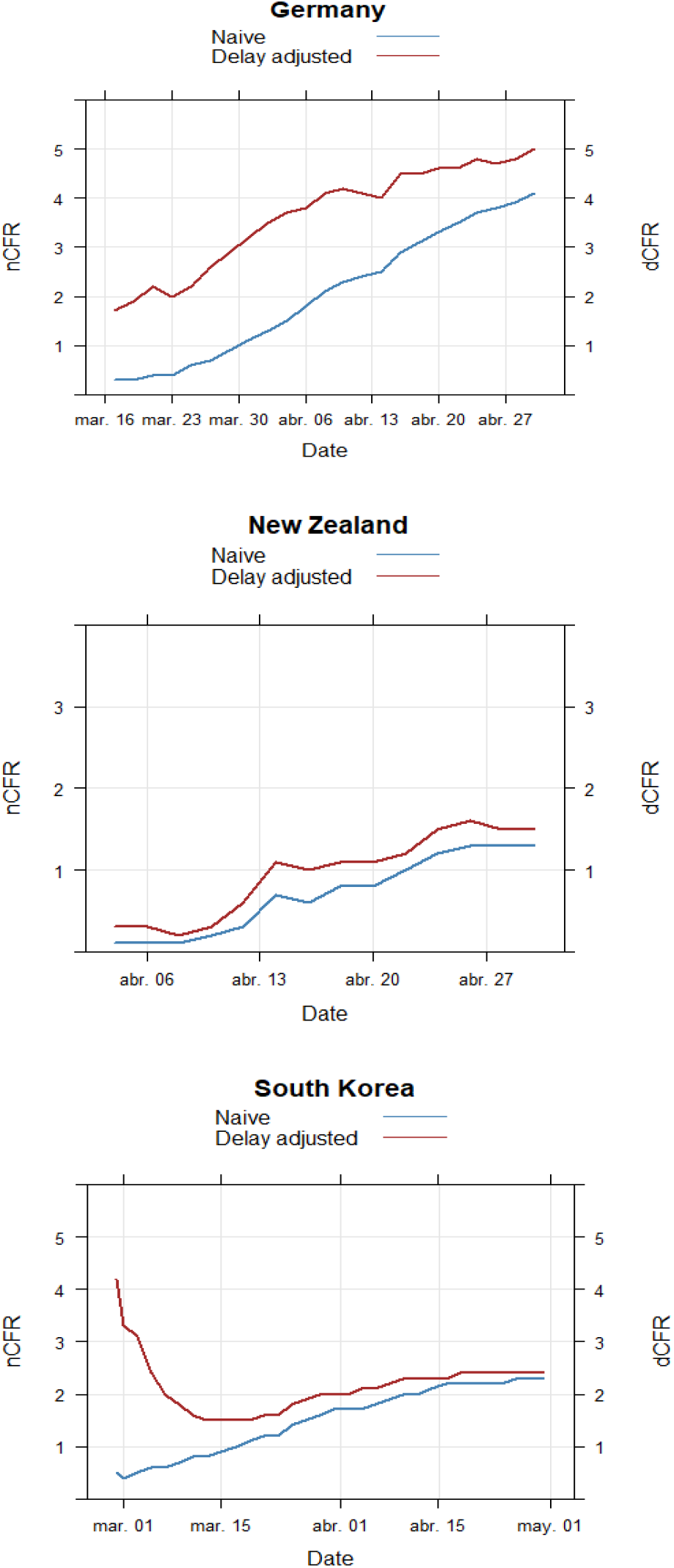
Fatality rate curve over time using the naive and the delay adjusted method for selected countries.

## 4. Discussion

The most remarkable result to emerge from the data is that our best approximation for the COVID-19 CFR across countries with a good testing coverage is of 2%. This estimation is an average of potentially different true CFR as they might vary depending on countries’ age structure (higher in countries with aged populations). Accordingly, this estimation based on the available data is lower than the reported by the WHO of around 3.4%.but still 20 times higher than the reported CFR for other diseases like seasonal influenza (0.01%) [21]. Moreover, it is lower than other recent coronavirus epidemics as the Severe Acute Respiratory Syndrome (SARS) with a fatality rate of 10% [22], or the Middle East Respiratory Syndrome (MERS) with an even higher mortality rate of around 34% [23]. We selected countries with a high testing coverage for our primary analysis to provide more robust results, however, further population seroprevalence studies will be required to estimate the infectious fatality rate which in simulation studies have been estimated to be lower than 1% [24].

Importantly, we have verified a positive correlation with the population age structure. Larger delay adjusted CFRs were observed in countries like Canada, Slovenia or Denmark which have a high proportion of people over 65 years old [12]. In addition, it have been reported that the outbreak specifically affected the older segment of the population in Canada, where more than 90% of who have died were over 60 years old, and nearly half of the deaths occurred in long term care homes [25], something that has been also reported in Slovenia [26]. On the other side, Singapore had the smallest CFR, partly due to the smaller proportion of older people (11%), and because the outbreak concentrated in younger migrant workers.

We showed that naïve CFR underestimates corrected estimates by a median of 1.3%, with the magnitude of this difference being correlated to the epidemic curve growth. This observation should be considered by policy makers when communicating the consequences of the outbreak. This bias, due to the calculation method, has been previously reported during the SARS epidemic, in which naïve fatality rate increased over time, leading some to conclude that was more lethal than it resulted to be. The public health impact of inaccurate estimates, resulting in misinformation and conflicting messages, can therefore exacerbate public alarm [7].

Some previous analysis has also attempted to estimate the CFR with different limitations that we have attempted to overcome. One study adjusted the delay time between diagnosis to death an reported CFR for 82 countries outside China of 4.24%, but this was calculated using a fairly simplistic method, based on the number of cases of 13 days previous to the assessment date [27]. Another study collected data from several surveillance sources of cases from mainland China only, adjusting for under-ascertainment, and time delay; the authors obtained a CFR of 1.38%, increasing to 6.4% in those older than 64 years [24]. The Centre of Evidence Based Medicine used a meta-analytic approach to calculate a prediction interval across countries of between 0.84% to 8.67%. The authors did not provide a pooled estimate due to the large heterogeneity observed. However, by using a fixed effect model the authors assumed the existence of an only true value, an assumption that is not appropriate to account for different true CFR across countries, they relied in naive estimates, and did not considering testing coverage in their assessment.

Recently, two non-peer reviewed seroprevalence studies reported estimates for the infectious fatality rate. In the first one, researchers from the university of Bonn took serological samples from approximately 1,000 inhabitants of the German town of Gangelt (population of 12,529 people), estimating an infection rate of 14%, and a fatality rate of 0.37%. Another study conducted in the Santa Clara County, California, United States, found a population-weighted prevalence that ranged from 2.5% to 4.2% and an infectious fatality rate of 0.12% to 0.2% [28]. However, these results have received some criticism related to the potential large false positive results from serology tests, and the recruitment method employed that might have overestimated the number of infected people.

We are aware that our estimates are based on the reported number of cases with a positive test result and not in the real number of infected people, overestimating the CFR. Asymptomatic cases and mild symptomatic patients are not routinely tested as it is underscored by recent publications. In a large-scale COVID-19 diagnostic testing in Iceland with 9,199 persons, found that 43% of positive cases were asymptomatic at the time of testing [22]. In another study, from a total of 215 pregnant women, 29 of 33 (88%) who were SARS-CoV-2 positive at admission were asymptomatic [29]. Although, the number of tests performed might differ from the number of individuals tested, and the distinction is not always clear in public data (i.e. Hong Kong reports in number of tests, while Singapore in number of people) [11], including countries with a large number of tests performed and low positive rate in our main analysis, has likely reduced the impact of the bias due to the ascertainment of cases.

We should also consider the bias due to underreporting of the cause of death, which goes in the opposite direction (underestimating CFR). For example, China has recently revised its death tolls, on April 17, adding 1,300 fatalities to its initial official count for the city of Wuhan; due to the inclusion of deaths that occurred at home or at institutions [30]. Similarly, in Madrid, Spain, around 4,100 death cases occurred among elder people in long term care homes who reported symptoms compatible with COVID-19 and that were not accounted in official reports [31]. One strategy to address this bias is comparing the all-cause mortality estimates with previous years, as in the Guayas region of Ecuador in which an excess of more than 6,000 deaths was observed, probably be related to the COVID-19 outbreak [32]. Based in this approach, approximately a 25% of the comparative excess mortality could be also attributed to COVID-19 in countries like Germany or Portugal [33].

We consider our calculations also have strengths, compared with previous estimation across countries, because: 1) we included countries with more than 500 confirmed cases which making calculations more stable, 2) we used an appropriate method for pooling proportions, including a random model that considers the different true estimates across different settings, 3) instead of using a naïve estimation, we applied a correction method to consider the delay time between diagnosis and death, and 4) we only included in our main analysis countries with good testing coverage, which might reduce the impact of the different biases due to ascertainment of cases.

### 4.1 Conclusion

In conclusion, assessing the fatality rate of COVID-19 is critical to determine the appropriateness of mitigation strategies, as well as to enable forecasting of healthcare requirements as the epidemic unfolds. Thus, one urgent need is to conduct large high quality seroprevalence studies [34]. Meanwhile, our findings contributes to the literature providing a good approximation of the true fatality rate across countries based in more appropriate methods and taking into account the testing coverage and positivity rate. Until more robust estimates are available, a CFR of 2% might be used for policy makers for comparability. Our study shows that naive estimation of CFR underestimates the potential threat of COVID-19, and although use of such methods are clearly easier to communicate to policy makers and the public, their use could be misleading for the deployment of health systems responses.

## Data Availability

Data used for the analysis is described in the supplementary file.

## Author contributions

**Carlos Canelo-Aybar, Jessica Beltran:** Conceptualization, **Carlos Canelo-Aybar, Jessica Beltran, Marilina Santero, Pablo Alonso-Coello:** Methodology; **Carlos Canelo-Aybar:** Data Curation and Analysis; **Carlos Canelo-Aybar, Jessica Beltran:** Writing-Original Draft; **Marilina Santero, Pablo Alonso-Coello:** Writing - Review & Editing; **Pablo Alonso-Coello:** Supervision.

## Funding

This research did not receive any specific grant from funding agencies in the public, commercial, or not-for-profit sectors.

## Conflict of interest

The authors declare that there are no conflicts of interest

## Notes

### Competing Interest Statement

The authors have declared no competing interest.

